# Technical and ethical challenges in polygenic embryo selection

**DOI:** 10.1101/2024.05.28.24308092

**Authors:** Shinichi Namba, Masato Akiyama, Haruka Hamanoue, Kazuto Kato, Minae Kawashima, Itaru Kushima, Koichi Matsuda, Masahiro Nakatochi, Soichi Ogishima, Kyuto Sonehara, Ken Suzuki, Atsushi Takata, Gen Tamiya, Chizu Tanikawa, Kenichi Yamamoto, Natsuko Yamamoto, The BioBank Japan Project, Norio Ozaki, Yukinori Okada

## Abstract

Whereas best practice of clinical prediction of human phenotypes by polygenic risk score (PRS) has yet to be fully implemented, commercial industries already offer pre-implantation genetic testing for PRS (PGT-P) to select embryos with ‘desirable’ potential. However, its efficacy is questionable due to the current technical challenges, which also raise ethical concerns. Our *in-silico* simulations utilizing biobank resources revealed that the embryo selected by PGT-P substantially differs depending on the choice of methods and the random fluctuation of the PRS construction. Here, we outline the technical challenges and also the ethical concerns that remain even if the technical challenges are solved, and hope to call on a society-wide discussion for this technology.

## Main

Estimating genetic susceptibility to clinical and non-clinical phenotypes has been increasingly successful with the advent of genome-wide association study (GWAS) and its derived score, polygenic risk score (PRS)^1^. PRS is generally calculated by aggregating the effects of common genetic variants associated with a phenotype and can effectively predict disease risk and other phenotypic values. Clinical implementation of PRS is one of the active research fields for the early detection and intervention of human diseases^2^. However, prior to clinical implementation, private enterprises marketed PRS analyses of embryos as pre-implantation genetic testing (PGT; PGT-P) for *in vitro* fertilization^3^. They claim that selected embryos have the potential to acquire desirable traits and are less susceptible to diseases. Although they are advancing ahead with PGT-P^4^, this technology raises various questions about its efficacy and ethical validity.

In this comment, we summarize the challenges and limitations of PRS which affects the efficacy of PGT-P, and highlight the non-deterministic nature of PRS by virtually selecting embryos using different PRS methods. We then address the ethical concerns of PGT-P and outline issues to be addressed and communicated society-wide.

### Current challenges and limitations of PRS in applying to PGT-P

One of the important challenges in using PRS for embryo selection is that PRS captures relatively limited phenotypic variance than consumers might expect. PRS explains no more than 5.8% of the variance in body mass index in Martin AR *et al*.,^5^ for example, and typically much less for diseases, often falling below 5% and even 1%^6^. The main reason for this is the difficulty in constructing PRS because the effect sizes of common variants are relatively small and estimated with error^7^. Due to this small variance captured by PRS, phenotypic gain by PGT-P will be inconspicuous and may not be clinically meaningful. The embryos from the same parents are genetically akin to each other just like dizygotic twins, making the expected phenotypic gain in PGT-P even smaller^8^. For example, the average gain was estimated to be 2.5 cm in height^9^, one of the most successful targets of PRS. The distribution of embryonic PRS will be even narrower than that in a general population. In addition, there are genetic determinants not captured by common variants (e.g., rare pathogenic variants), which can have large effect sizes at the individual level. The effect sizes of rare variants were indeed estimated to be up to 1.8cm for height in the Japanese population^10^, which is comparable to the gain by PGT-P. While one company has reported its methodology for genotyping both common and rare variants^11^, the way to integrate them into a calibrated score has yet to be established. Therefore, it will be difficult to justify the efficacy of PGT-P postnatally, and PGT-P may be disappointing for parents wishing to receive PGT-P.

PRS might have unintentional effects on non-targeted phenotypes because one variant can be associated with multiple phenotypes (i.e., pleiotropy). Epidemiological disease correlations can also cause unintentional pleiotropic effects. It is difficult to forecast the pleiotropy, or more generally, to reveal the biology of PRS because most variants used for PRS are not causal by themselves, merely tagging correlated causal variants.

Moreover, the characteristics of PRS depend on the discovery GWAS from which PRS was constructed. Sample size and subsequent statistical power of the GWAS determine the maximum accuracy of phenotypic prediction by PRS. In addition, if GWAS is confounded by factors other than direct phenotypic relationships, PRS would be confounded likewise. As the confounding factors include ethically controversial and ancestry-correlated traits like skin tanning and hair color^12^, selecting embryos based on a particular phenotype may actually result in selecting particular ancestries. In addition, PRS distribution can be implausibly different across populations^13^, which might also lead to the selection of particular ancestries when parents’ populations are different. Finally, PRS is less predictable when applied to populations not mainly included in its discovery GWAS. As GWAS has been conducted primarily on the European population, PRS predictability is generally lower in other underrepresented populations, which may foster health disparities^5^.

### Selected embryos vary across PRS construction methods

Various PRS construction methods have been proposed^1,14–18^ to improve the accuracy of phenotype prediction by PRS. While these methods are gaining popularity in PRS studies, no single state-of-the-art method exists, and individual researchers pick up their preferred method. Through our simulation utilizing biobank resources, we showed that this concern is indeed one of the biggest problems with PGT-P.

First, we constructed PRS for adult height using six popular PRS methods^1,14–18^. By using publicly available genotype data from Biobank Japan, we virtually created a random mate pair and generated genotypes of ten embryos by simulating recombination between the haplotypes of the mate pair. We then ranked the embryos in the descending order of PRS (**Supplementary Methods**). We repeated this process 500 times and in no more than half the cases was the same embryo selected as the top-ranked one in any combinations of PRS methods (median 30.0% [range, 20.4–41.6%], **Figure 1a**). Strikingly, the top-ranked embryo by a particular PRS method was lowest-ranked by at least one other method with a median 5.9% chance (3.4–8.0%, **Figure 1b**), and vice versa (median 5.2% [4.0–7.8%], **Figure 1c**). These results imply that unselected embryos would be selected if other PRS methods were used, raising serious ethical concerns about selecting embryos and consequently discarding other embryos in an unreliable way.

**Figure 1.**
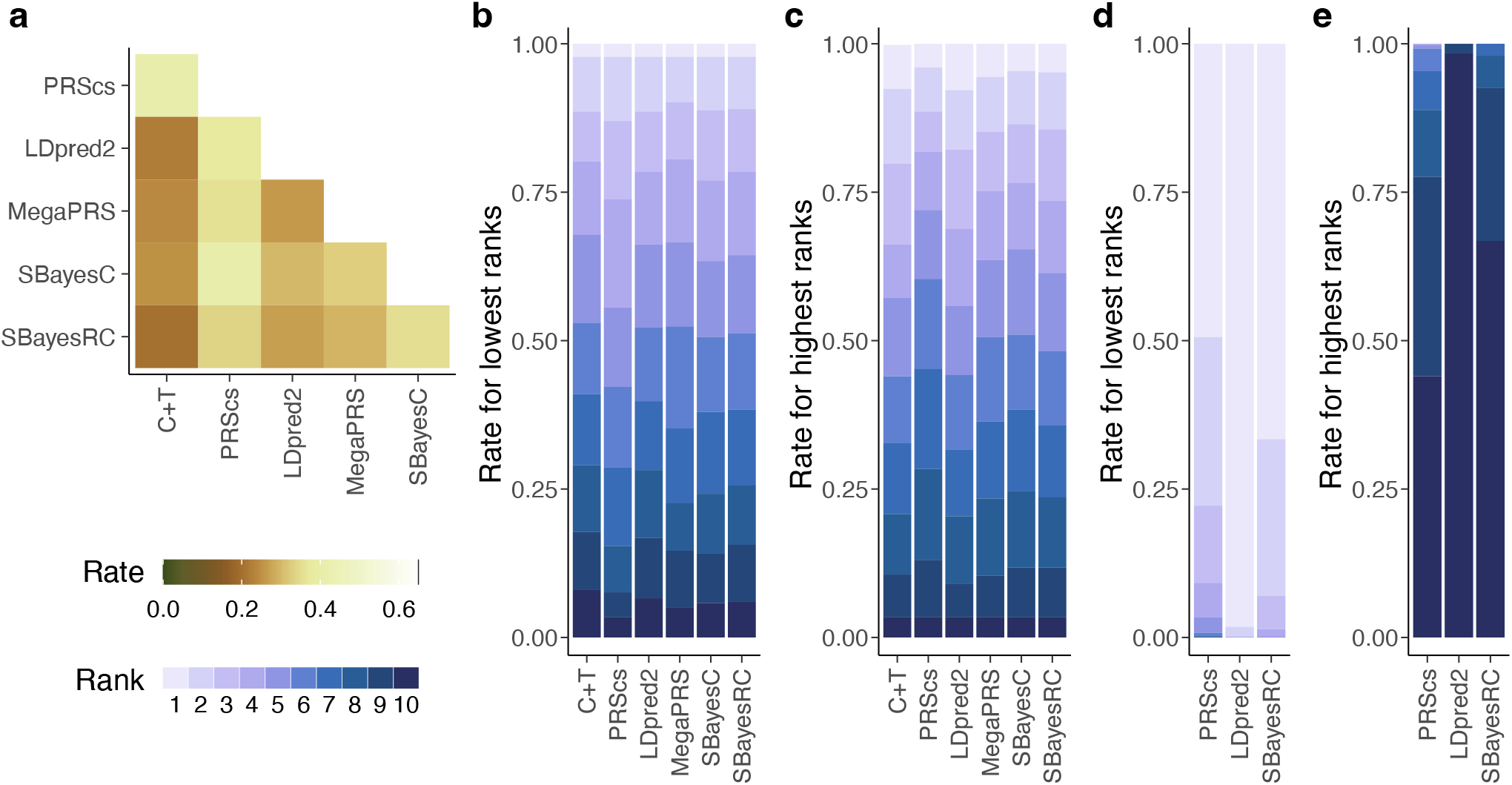
Prominent inconsistency of embryo selection rank across PRS methods. **a**, the rate at which two PRS methods chose the same embryo as the top-ranked in 500 simulations. **b**, the distribution of lowest ranks across all PRS methods for the embryo top-ranked by the PRS method in the *x* coordinate. Rank 1 means the embryo top-ranked by the PRS method in the *x* coordinate was also top-ranked by any other methods. On the contrary, rank 10 means the embryo top-ranked by the PRS method in the *x* coordinate was bottom-ranked by at least one other PRS method. **c**, same as **b**, but of the highest ranks for the bottom-ranked embryo. **d**, the distribution of lowest ranks among 10 PRS replicates by the same method, for the embryo top-ranked by the PRS constructed in the first repeat. **e**, same as **d**, but of the highest ranks for the bottom-ranked embryo. PRS, polygenic risk score. C+T, clumping and thresholding.

As a larger GWAS sample size improves accuracy in phenotype prediction by PRS, one might think that a larger GWAS sample size might also solve the inconsistent selection of embryos between PRS methods. We simulated saturated GWAS summary statistics^19^ with a sample size larger than any GWAS to date (*N*_*case*_ = 5×10^6^ and *N*_*control*_ = 5×10^6^) and with high heritability (*h*^*2*^ = 0.8), and repeated the embryo ranking simulation (**Supplementary Methods**). However, the top-ranked embryos were still different across PRS methods at a rate similar to the previous one (median 24.0 [18.6-34.8%], **Supplementary Figure 1**), indicating that the inconsistency between PRS methods cannot be easily solved in the current GWAS framework.

The inconsistent selection of embryos highlights the difference in statistical models across PRS methods. They use different formulas and make different assumptions for the distribution of causal variants in the human genome^1,14–18^. As some PRS methods adopt Bayesian optimization, even simply repeating the same PRS method produced a slightly different embryo ranking (**Figure 1d and e**). This non-deterministic nature of PRS illustrates that PRS construction is a probabilistic process with fluctuation, and makes PGT-P distinct from other PGTs. Although there is an established PGT targeting genetic variants (PGT for monogenic diseases, PGT-M), its targets are rare variants with verified pathogenicity. In contrast, embryos with a high score of a disease PRS is merely predicted to be at high risk by a particular statistical model with random fluctuation.

### Ethical concerns of PGT-P independent of PRS efficacy

As shown in the simulations, embryo selection by PRS is currently not robust enough to be implemented as PGT-P. Moreover, the concept of PGT-P itself may raise yet other ethical concerns. PGT-P sorts out the value of embryos along with arbitrary traits desirable for consumers, leading to potential eugenic thinking. Although the established PGTs other than PGT-P also sort out embryos based on their genetic features, their purposes are treating infertility and preventing severe genetic disorders, different from PGT-P. Even for these PGTs, each country has set law and ethical guidelines after long debates. In Japan, as in some countries^20^, the target diseases of the established PGTs are strictly limited, and the PGTs are not practiced for screening purposes. Only parents approved by specialists’ review are able to receive PGTs after repeated genetic counseling^21^. Therefore, there is a gap between the current ethical guidelines and the targeting of healthy embryos by PGT-P. Considering the strict restrictions of the target diseases for the established PGTs, it is also questionable whether common diseases should be subject to embryo selection because common diseases include to some extent preventable and controllable diseases by the postnatal lifestyle.

Another ethical concern is the PGT-P’s effects on children’s identity establishment. When consumers have excessive expectations for the effectiveness of PGT-P, PGT-P might negatively affect the parent–child relationship. For example, children may feel burdened that they were selected because they are ‘genetically superior’ if informed that they were born with PGT-P. Besides these concerns about increasing power imbalances between parents and children, the impact of unexpected changes in the environment of children born with PGT-P, such as the death or remarriage of their parents, is also unknown. Genetic counseling should be at least mandatory to accurately understand the risk-benefit of PGT-P. Nonetheless, counseling parents alone is insufficient, as it does not necessarily convey accurate knowledge to their children. As there already exist children born with PGT-P, careful support for these children is also required. At the same time, it is important not to treat children differently for being born PGT-P in order to respect their dignity.

Despite these ethical concerns, more than half of the participants in a U.S. survey reported no moral objection to PGT-P and the participants showed a willingness to use PGT-P to some extent^22^. Because this survey was taken place in the U.S., a country where PGT-P is already provided as a private service, we might not jump to generalize this survey report globally. However, it is safe to say that the experts may not have communicated the ethical concerns to the public appropriately, while PGT-P seems to be in demand. Discussing the technical challenges and the ethical concerns by both experts and the public is mandatory to regulate PGT-P or seek its tolerable usage in preparation for future technical innovations.

## Conclusion

We summarize current technical challenges and ethical concerns for PGT-P in **Box 1**. PGT-P to date is not sufficiently effective or robust for embryo selection. Technical challenges stemming from PRS also need to be addressed, and these challenges are generally applicable to the clinical implementation of PRS. Some ethical concerns arise from the technical challenges, including the unreliable and confounded selection of embryos. In this regard, we provide a new viewpoint by showing the inconsistent selection of embryos across PRS construction methods and across replicates of the same methods. Furthermore, there are ethical concerns about sorting out life and children’s self-identity that technical development cannot solve. These concerns must be addressed before PGT-P is accepted as an ethically approved examination. Given these challenges and concerns, we advocate that PGT-P is currently premature for implementation, although we take seriously the desire of parents to take on the new challenge of selecting embryos using PGT-P for a variety of reasons. We clarify that these concerns do not affect the dignity of children already born with PGT-P. We rather call on a society-wide discussion and expect it will be consistent with previous arguments on the established PGTs.

## Figure and box descriptions

**Box 1. Technical challenges and ethical concerns in PGT-P**.

- Technical challenges specific to PGT-P
  ➢ The expected phenotypic gain by PGT-P can be too small to be validated or to have clinical meanings.
  ➢ The selected embryo differs across PRS construction methods and even across replications of the same method.
- Technical challenges stemming from PRS
  ➢ PRS ignores genetic determinants other than common variants.
  ➢ PRS can have unintentional pleiotropic effects on non-targeted phenotypes.
  ➢ PRS can take over the confounding factors of its discovery GWAS.
  ➢ The efficacy and distribution of PRS are different across populations.
- Ethical concerns
  ➢ PGT-P may be used eugenically to sort out the value of life.
  ➢ Without appropriate support, children born with PGT-P may have difficulty establishing their identity.
  ➢ Public understanding of ethical concerns is not sufficient.

## Data Availability

The genotype data of Biobank Japan are available at NBDC Human Database (https://humandbs.biosciencedbc.jp/en/) with the accession ID of JGAS000114 and JGAS000412. The GWAS summary statistics for height and the height PRS calculated with SBayesC are publicly available at https://portals.broadinstitute.org/collaboration/giant/index.php/GIANT_consortium_data_files.

## Data and code availability

The genotype data of Biobank Japan are available at NBDC Human Database (https://humandbs.biosciencedbc.jp/en/) with the accession ID of JGAS000114 and JGAS000412. The GWAS summary statistics for height and the height PRS calculated with SBayesC are publicly available at https://portals.broadinstitute.org/collaboration/giant/index.php/GIANT_consortium_data_file s. We constructed PRS using the following publicly available tools: LDpred2 (https://privefl.github.io/bigsnpr/articles/LDpred2.html), PRScs (https://github.com/getian107/PRScs), SBayesC (https://cnsgenomics.com/software/gctb), SBayesRC (https://github.com/zhilizheng/SBayesRC), and MegaPRS (https://dougspeed.com/).

## Acknowledgment

S.N. was supported by Takeda Science Foundation. Y.O. was supported by JSPS KAKENHI (22H00476), AMED (JP21gm4010006, JP22km0405211, JP22ek0410075, JP22km0405217, JP22ek0109594, JP223fa627002, JP223fa627010, JP233fa627011, JP23zf0127008), JST Moonshot R&D (JPMJMS2021, JPMJMS2024), Takeda Science Foundation, Bioinformatics Initiative of Osaka University Graduate School of Medicine, Institute for Open and Transdisciplinary Research Initiatives and Center for Infectious Disease Education and Research (CiDER), and Center for Advanced Modality and DDS (CAMaD), Osaka University. N.O. was supported by AMED (JP22dk0307113).

## References

1. Choi, S. W., Mak, T. S.-H. & O’Reilly, P. F. Nat. Protoc. 15, 2759–2772 (2020).

2. Torkamani, A., Wineinger, N. E. & Topol, E. J. Nat. Rev. Genet. 19, 581–590 (2018).

3. Goldberg, C. Bloomberg https://www.bloomberg.com/news/articles/2021-09-17/picking-embryos-with-best-health-odds-sparks-new-dna-debate (2021).

4. Kumar, A. et al. N. Engl. J. Med. 385, 1726–1727 (2021).

5. Martin, A. R. et al. Nat. Genet. 51, 584–591 (2019).

6. Lambert, S. A. et al. Nat. Genet. 53, 420–425 (2021).

7. Vilhjálmsson, B. J. et al. Am. J. Hum. Genet. 97, 576–592 (2015).

8. Turley, P. et al. N. Engl. J. Med. 385, 78–86 (2021).

9. Karavani, E. et al. Cell 179, 1424-1435.e8 (2019).

10. Akiyama, M. et al. Nat. Commun. 10, 4393 (2019).

11. Kumar, A. et al. Nat. Med. 28, 513–516 (2022).

12. Sanderson, E., Richardson, T. G., Hemani, G. & Davey Smith, G. Int. J. Epidemiol. 50, 1350–1361 (2021).

13. Martin, A. R. et al. Am. J. Hum. Genet. 100, 635–649 (2017).

14. Privé, F., Arbel, J. & Vilhjálmsson, B. J. Bioinformatics 36, 5424–5431 (2021).

15. Ge, T., Chen, C. Y., Ni, Y., Feng, Y. C. A. & Smoller, J. W. Nat. Commun. 10, 1–10 (2019).

16. Lloyd-Jones, L. R. et al. Nat. Commun. 10, 5086 (2019).

17. Zheng, Z. et al. bioRxiv (2022) doi:10.1101/2022.10.12.510418.

18. Zhang, Q., Privé, F., Vilhjálmsson, B. & Speed, D. Nat. Commun. 12, 4192 (2021).

19. Yengo, L. et al. Nature 610, 704–712 (2022).

20. European Society of Human Reproduction and Embryology. https://www.eshre.eu/-/media/sitecore-files/Press-room/Resources/2-Regulation.pdf (2017).

21. Sugiura-Ogasawara, M. & Sato, T. Nat. Med. 28, 1732–1733 (2022).

22. Meyer, M. N., Tan, T., Benjamin, D. J., Laibson, D. & Turley, P. Science 379, 541–543 (2023).

