## Supplementary Information for "Technical and ethical challenges in polygenic embryo selection"

S Namba *et al.*

Corresponding to Shinichi Namba

and Yukinori Okada

### Supplementary Figures

**Supplementary Figure 1. Prominent inconsistency of embryo selection rank across PRS methods using a large-scale case–control simulated GWAS summary statistics.**

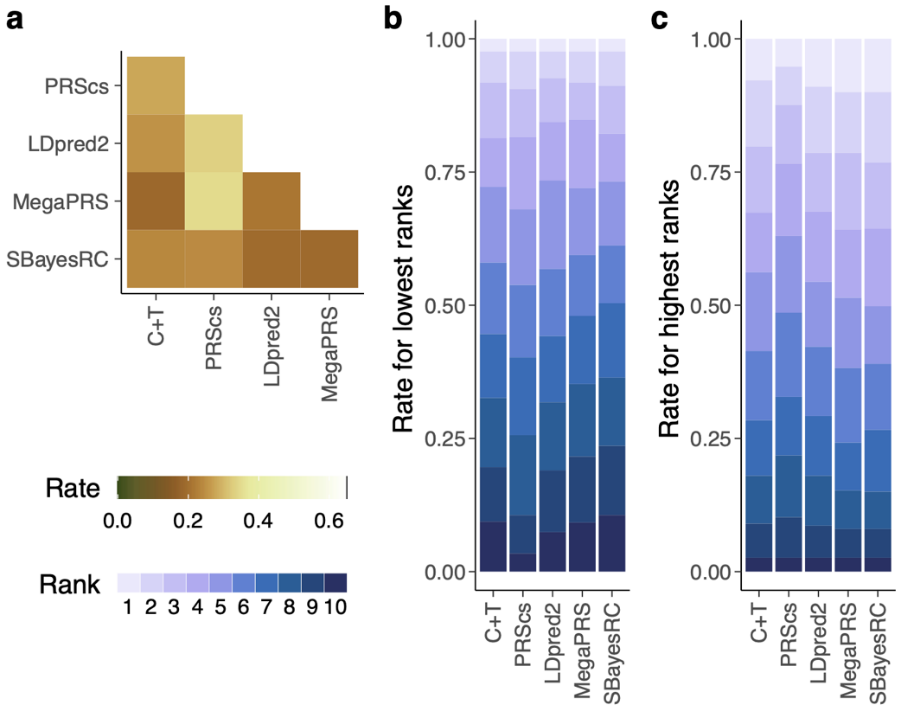

Same as **Figure 1**, but using simulated summary statistics for PRS construction. **a**, the rate at which two PRS methods chose the same embryo as the top-ranked in 500 simulations. **b**, the distribution of lowest ranks across all PRS methods for the embryo top-ranked by the PRS method in the *x* coordinate. **c**, same as **b**, but of the highest ranks for the bottom-ranked embryo. We excluded SBayesC because we observed a convergence issue and found markedly lower correlations with PRS calculated with other methods. PRS, polygenic risk score. C+T, clumping and thresholding.

### Supplementary Methods

#### Biobank Japan

Biobank Japan is a hospital-based cohort of approximately 270,000 participants, all of whom were diagnosed with at least one of the main target diseases of Biobank Japan[1]. All the participants provided written, informed consent approved by ethics committees of the Institute of Medical Sciences, the University of Tokyo and RIKEN Center for Integrative Medical Sciences. Although a large subset of Biobank Japan (169,009 participants) was included in the discovery cohorts of the height GWAS[2], we used the remaining participants for the embryo simulation. The participants were genotyped with the Illumina Infinium Asian Screening Array. We excluded individuals with a low call rate (<0.98) and outliers from the East Asian cluster based on a principal component analysis. We excluded the variants meeting the following criteria: (i) with a low call rate (<0.99); (ii) with low minor allele counts (<5); and (iii) with Hardy–Weinberg equilibrium test $P$ values $< 1.0 \times 10^{-10}$. We statistically phased the genotype data using Shapeit4[3] and performed imputation with the combined reference panel of 1000 Genomes Project Phase 3 and whole-genome sequencing data of 1,037 Japanese samples[2] using Minimac4[4]. After imputation, we retained variants with MAF larger than 0.01 and an info score larger than 0.7. We retained the participants in the Japanese Hondo (i.e., main islands) cluster by visual inspection of principal component analysis. We used king[5] to exclude relatives within 2 degrees. Out of the 50,212 samples that passed the quality controls, we randomly selected and matched 1,000 samples to generate 500 virtual mate pairs for embryo simulation.

#### Simulation of embryo genotypes

We simulated embryo genotypes by modeling recombination as a Poisson process. Briefly, we obtained genetic distance in cM from the HapMap genetic map and randomly assigned the positions of crossovers so that the number of crossovers matched the expectation from the genetic distance. We leveraged an established code of embryo simulation[6] (https://bitbucket.org/ehudk/nembryo-pgs-selection/src/master/) to simulate ten embryo genotypes per mate pair.

**Polygenic risk scores (PRS)**

We calculated PRS with six PRS construction methods, Clumping and Thresholding (C+T), LDpred2-auto[7], PRScs-auto[8], SbayesC[9], SbayesRC[10], and MegaPRS[11]. These PRS construction methods use GWAS summary statistics and a linkage disequilibrium (LD) reference panel and do not require a tuning/validation cohort. The details of respective PRS construction methods are described below. We used the summary statistics from an East Asian-specific GWAS meta-analysis for height[12] ($N$ = 363,856) and a simulated case–control GWAS summary statistics. For the LD reference panel, we used East Asian participants ($N$ = 504) of the 1000 Genomes Project Phase 3 to facilitate the reproducibility of our PRS construction.

Clumping and Thresholding (C+T)

C+T is a simple PRS construction method that aggregates raw effect sizes of selected variants. We retained variants with $P < 5\times10^{-8}$ and performed LD clumping using plink v1.90b6.16[13] with the option "plink --clump --clump-kb 250 --clump-r2 0.1".

LDpred2-auto

LDpred2, as well as the other PRS methods used here except for C+T, is a Bayesian approach that infers variants' posterior mean effect sizes[7]. LDpred2 assumes a non-infinitesimal distribution of effect sizes and uses a point-normal mixture prior. LDpred2-auto is a mode of LDpred2 that automatically estimates the hyperparameters of LDpred2, specifically, heritability and sparsity of causal variants. We used LDpred2 implemented in the bigsnpr R package v1.11.6 with the options recommended by the authors (https://privefl.github.io/bigsnpr/articles/LDpred2.html). Specifically, we used the snp_ldpred2_auto() function with the "vec_p_init = seq_log(1e-4, 0.2, length.out = 30), allow_jump_sign = FALSE, shrink_corr = 0.95" option. We also specified the heritability estimated by the LD score regression[14] reimplemented in the bigsnpr R package as the heritability used for initialization. For the reference panel, we used the HapMap3+ variants, an extended set of approximately 1.4 million variants with better genome coverage than the HapMap3 variants[15], of the East Asian participants of the 1000 Genomes Project phase 3.

PRScs-auto

PRScs is a Bayesian regression framework using continuous shrinkage priors to improve computational efficiency and accurately model local LD patterns[8]. In the default mode (i.e., PRScs-auto), the global shrinkage hyperparameter is automatically learned from GWAS summary statistics itself. We ran PRScs (version Apr 6, 2021) with the accompanying reference panel derived from the HapMap3 variants of the East Asian participants of the 1000 Genomes Project phase 3.

SBayesC

SBayesC is a Bayesian approach using the same point-normal mixture prior as used in the LDpred2 method[9]. The height PRS for the East Asian population was calculated in the original GWAS article using SBayesC and is available publicly[12]. We note that the GWAS used for the SBayesC PRS included additional samples from 23andMe (*N*=472,730 in total), which were not included in the publicly available GWAS summary statistics used for the other PRS methods. We used SBayesC implemented in the GCTB software (v2.03 beta) for the simulated GWAS summary statistics; however, we observed a convergence issue and found markedly lower correlations with PRS calculated with other methods. Therefore, we excluded the SBayesC PRS from the analyses using the simulated GWAS summary statistics.

SBayesRC

SBayesRC is an extension of SBayesR[12], a Bayesian approach that models variant effect sizes using a flexible mixture of a point-mass Dirac distribution on zero and multiple Gaussian distributions with different variances[10]. SBayesRC incorporated functional annotations (specifically, general annotations curated by the BaselineLD v2.2 model[16]) to modulate the probability that every variant belongs to the individual effect size distributions. We used the SBayesRC R package v0.1.4 and the accompanied LD reference panel derived from the East Asian participants of the UK Biobank.

MegaPRS

MegaPRS is a suite of PRS methods that were reimplemented to model variant effect sizes with the BLD-LDAK heritability model, where variant effect sizes depended on minor allele frequency (MAF), local levels of linkage disequilibrium, and function annotations[11]. Among the PRS methods available in MegaPRS, the LDAK-BayesR-SS method, an implementation of SBayesR, was reported to produce the most accurate PRS[11] and set as the software default. We used MegaPRS implemented in LDAK v5.2 and followed the authors' recommendation for its options (https://dougspeed.com/). First, we estimated heritability partitioned by functional annotations with the BLD-LDAK model. Then, we applied the LDAK-BayesR-SS method with the "--cv-proportion .1 –window-cm 1 –allow-ambiguous YES" option and specified the accompanying high LD region data.

**Simulation of case–control GWAS summary statistics**

We started with the haplotype data of the East Asian participants of the 1000 Genomes Project Phase 3. We restricted the variants used to simulate GWAS summary statistics to those included in the height GWAS summary statistics and available in the Biobank Japan data. We partitioned the genome into the 1,445 non-overlapping LD blocks as reported previously[17] and randomly chose one to three variants with MAF larger than 0.01 as causal variants per LD block. Consequently, we assigned 2,875 variants as causal in total. Following the GCTA model[18], we simulated causal effect sizes assuming that the causal effect sizes of standardized genotypes were independent from each other and followed the Gaussian distribution with a mean of zero. We scaled the causal effect sizes so that the variance explained aggregated across all causal variants (i.e., heritability) matched to 0.8. We then used the simGWAS R package v0.2.0-4[19] per LD block to simulate case–control GWAS summary statistics with a large sample size ($N_{case} = N_{control} = 5\times10^6$). Unlike other GWAS simulation tools, simGWAS skips assigning individual phenotypic values and directly simulate GWAS summary statistics with any sample size, although it supports only case–control GWAS simulation. We confirmed that most of the causal variants were significant ($P < 5\times10^{-8}$) in the simulated GWAS summary statistics (2,405 / 2,875 = 83.7%).

### Supplementary References

1. Nagai, A. *et al. J. Epidemiol.* **27**, S2–S8 (2017).
2. Akiyama, M. *et al. Nat. Commun.* **10**, 4393 (2019).
3. Delaneau, O., Zagury, J.-F., Robinson, M. R., Marchini, J. L. & Dermitzakis, E. T. *Nat. Commun.* **10**, 5436 (2019).
4. Das, S. *et al. Nat. Genet.* **48**, 1284–1287 (2016).
5. Manichaikul, A. *et al. Bioinformatics* **26**, 2867–2873 (2010).
6. Karavani, E. *et al. Cell* **179**, 1424-1435.e8 (2019).
7. Privé, F., Arbel, J. & Vilhjálmsson, B. J. *Bioinformatics* **36**, 5424–5431 (2021).
8. Ge, T., Chen, C. Y., Ni, Y., Feng, Y. C. A. & Smoller, J. W. *Nat. Commun.* **10**, 1–10 (2019).
9. Lloyd-Jones, L. R. *et al. Nat. Commun.* **10**, 5086 (2019).
10. Zheng, Z. *et al. bioRxiv* (2022) doi:10.1101/2022.10.12.510418.
11. Zhang, Q., Privé, F., Vilhjálmsson, B. & Speed, D. *Nat. Commun.* **12**, 4192 (2021).
12. Yengo, L. *et al. Nature* **610**, 704–712 (2022).
13. Chang, C. C. *et al. Gigascience* **4**, 7 (2015).
14. Bulik-Sullivan, B. *et al. Nat. Genet.* **47**, 291–295 (2015).
15. Privé, F., Albiñana, C., Arbel, J., Pasaniuc, B. & Vilhjálmsson, B. J. *bioRxiv* (2023) doi:10.1101/2022.10.10.511629.
16. Gazal, S. *et al. Nat. Genet.* **49**, 1421–1427 (2017).
17. Berisa, T. & Pickrell, J. K. *Bioinformatics* **32**, 283–285 (2016).
18. Yang, J., Lee, S. H., Goddard, M. E. & Visscher, P. M. *Am. J. Hum. Genet.* **88**, 76–82 (2011).
19. Fortune, M. D. & Wallace, C. *Bioinformatics* **35**, 1901–1906 (2019).
